# Image brightness indicates lumen direction in a patient-derived virtual colon: a learning-free approach to semi-autonomous colonoscopy

**DOI:** 10.64898/2026.08.31.26361807

**Authors:** Koji Kojima

## Abstract

**Background:** Automated colonoscope navigation requires a way to determine where the lumen continues. One possible approach is to estimate the distance from the endoscope tip to the surrounding wall. However, adding a range sensor to a conventional colonoscope is technically difficult, and depth estimation from monocular endoscopic images remains challenging. We considered a simpler possibility. Because the light source of an endoscope is located close to the camera, distant regions of the lumen tend to appear darker than nearby walls.

**Aim:** To determine whether lumen direction can be estimated from image brightness alone in a patient-derived virtual colon, and to compare this estimate with one obtained from true wall distances.

**Methods:** A virtual colon was reconstructed from the author’s CT colonography data (0.5 mm isotropic resolution). Endoluminal images were rendered at 342 positions along 1368 mm of the colonic centreline. At each position, the image was divided into 24 × 24 directional cells. Lumen direction was estimated from the darkness-weighted centroid of the largest connected component among the darkest 10% of cells. For comparison, the true distance from the virtual endoscope to the wall was obtained by ray casting, and a distance-based direction was calculated using the same method. Two illumination models were tested: a directional light without distance attenuation and a point source at the endoscope tip with distance attenuation.

**Results:** With point-source illumination, the brightness-based direction closely matched the distance-based direction, with a median angular difference of 2.5°. Differences greater than 30° occurred in 2.3% of frames. Within each frame, brightness and true wall distance were negatively correlated in all 342 frames (median Spearman ρ = −0.770).

Using the direction toward the centreline point 40 mm ahead as a fixed geometric reference, the median angular error was 6.6° for the brightness-based estimator and 6.5° for the distance-based estimator. The median frame-by-frame difference in error was −0.09°. In contrast, the brightness cue failed under directional illumination without distance attenuation (median difference from the distance-based direction, 43.1°).

**Conclusion:** In this patient-derived virtual colon, image brightness provided nearly the same lumen-direction information as exact wall distance when a point-source illumination model with distance attenuation was used. Exact metric distance was therefore not necessary for this specific navigation task. These findings suggest a simple, learning-free approach to semi-autonomous colonoscope steering based on image brightness and deterministic calculation.

## 1. Introduction

Automated colonoscope navigation requires the system to determine which direction the instrument should follow. Several approaches are possible. A system may learn an appropriate action from training data [1,2], or it may estimate the three-dimensional geometry of the lumen and calculate a path [3,4,5].

Both approaches can become technically complex. Machine-learning systems require appropriate training data and validation [1,2]. Geometry-based navigation requires information about the distance between the endoscope and the surrounding wall. Additional range sensors such as time-of-flight or structured-light devices may be difficult to incorporate into the small distal tip of a conventional colonoscope. Estimating depth from ordinary monocular endoscopic images is also difficult because the colonic wall has limited texture, strong reflections, fluid, deformation, and continuous changes in illumination [3,4,5].

However, an ordinary endoscopic image already contains a simple physical cue that may be useful for navigation: **brightness**.

The endoscope illuminates the same field that it observes. Nearby walls are generally strongly illuminated, whereas distant parts of the lumen receive less light and therefore tend to appear darker. This relationship was recognised in the earliest work on endoscope navigation, in which the dark region of the image was used directly as a navigational landmark [6,7,8,9,10]. Surface orientation also affects brightness [11]. Consequently, brightness cannot be used as a direct measurement of wall distance. Nevertheless, exact distance may not be necessary for navigation. A navigation system may only need to determine **which direction is more open**.

Experienced endoscopists routinely use this visual information. A dark region often indicates the direction in which the lumen continues. The important question is whether this intuitive cue is sufficiently reliable to be used computationally. Image-based steering systems built on this and related cues have reached in vitro and early clinical evaluation [12,13,14,15,16], but the accuracy of the brightness cue itself has not, to our knowledge, been measured against exact geometry.

This question is difficult to answer in a patient because the true distance from the endoscope to every part of the colonic wall is unknown. A virtual colon reconstructed from patient CT data provides a useful experimental environment because both the rendered endoscopic image and the exact three-dimensional geometry are available.

We therefore constructed a patient-derived virtual colon and asked a simple question:

**How closely does the lumen direction estimated from image brightness agree with the direction estimated from true wall distance?**

The purpose of this study was not to reconstruct three-dimensional geometry from brightness. Rather, we tested whether brightness alone provides enough directional information for a simple, calculation-based navigation system.

## 2. Methods

### 2.1 Virtual colon

The virtual colon was constructed from the author’s own CT colonography data [17]. The CT dataset had an isotropic resolution of 0.5 mm and was acquired in the supine position.

The colonic lumen was segmented in 3D Slicer using an attenuation range of −1000 to −400 HU. Surface smoothing was deliberately disabled to preserve haustral folds and fold convergence. Morphological closing was used to bridge gaps in the segmentation. Kernel sizes k3, k4, and k5 were tested, and k5 was selected because it provided the best luminal continuity.

The resulting surface contained 911,548 vertices and 1,823,284 triangles and had a bounding extent of 292.9 × 210.0 × 383.8 mm.

A luminal centreline was extracted using seeded flood fill from a surface-normal-offset seed point. The initial path length was 1606 mm. The terminal 160 mm, which entered the appendix and terminal ileum, was excluded. A total of 1368 mm of colon was analysed.

### 2.2 Rendering and illumination

Endoluminal images were rendered using VTK/PyVista with a vertical field of view of 120°. The colonic surface was assigned a mucosa-like diffuse material with specular reflection of 0.30, specular power of 18, and ambient illumination of 0.12.

Two illumination models were compared (Figure 1a).

**Figure 1.**
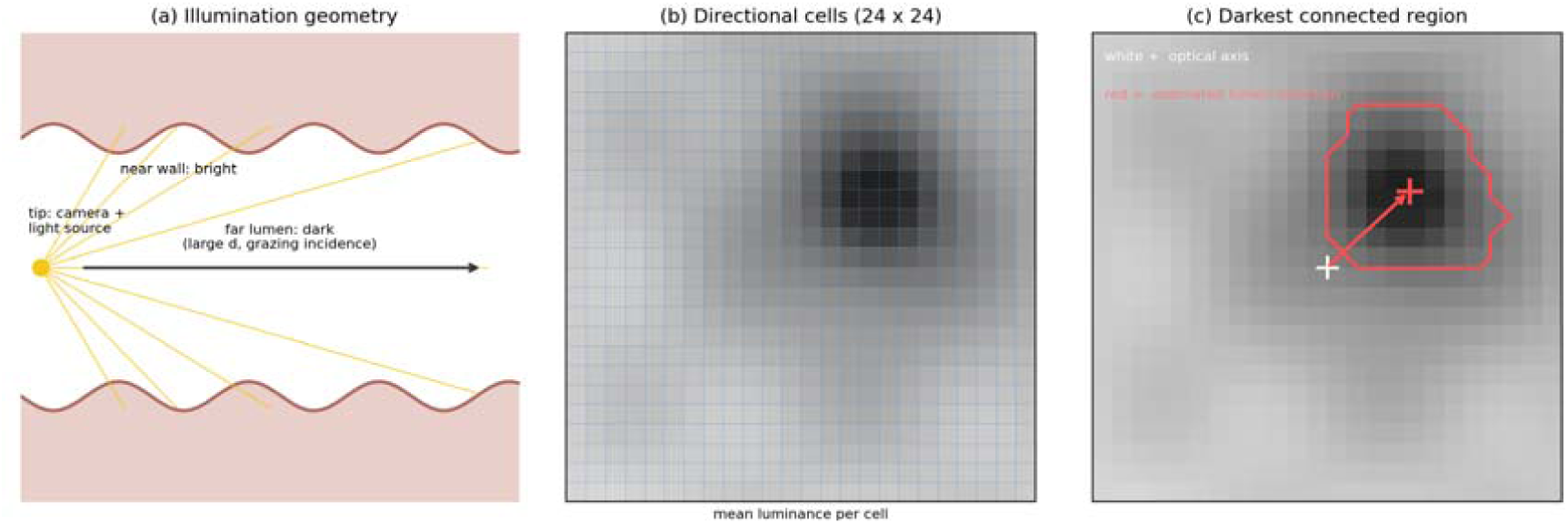
Principle and estimation procedure. (a) The endoscope illuminates the field that it observes. Because the source is co-located with the camera, radiance falls with both distance and incidence angle, and in a tubular organ both terms act in the same direction: the far lumen is dark, the near wall is bright. (b) The central portion of the rendered image is divided into 24 × 24 directional cells, and the mean luminance of each cell is computed. (c) The darkest 10% of cells are thresholded, the largest 4-connected component is retained, and its darkness-weighted centroid defines the estimated lumen direction. Panels (b) and (c) are schematic.

#### Directional illumination

A conventional directional headlight without distance attenuation was used. The illumination vector was constant across the image and brightness did not decrease with distance.

#### Point-source illumination

A positional light was placed at the virtual endoscope tip. Light intensity decreased with distance according to

1 / (1 + (d / 25 mm)^2^)

where d is the distance from the light source.

The two conditions differed only in the illumination model. The geometry, camera position, and distance calculations were unchanged.

### 2.3 Estimation of lumen direction

The virtual camera was placed at 342 positions along the centreline at intervals of 4 mm. At each position, the camera was directed toward the centreline point 12 mm ahead.

The outer 15% of the image was excluded, and the remaining image was divided into 24 × 24 cells (Figure 1b). Each cell represented a known direction in camera coordinates.

Mean luminance was calculated for each cell as 0.2126 R + 0.7152 G + 0.0722 B

Three methods were examined.

**Argmin:** the direction corresponding to the single darkest cell.

**Blob:** the darkest 10% of cells were identified. The largest 4-connected dark region was selected, and its darkness-weighted centroid was calculated (Figure 1c). This was the primary estimator.

**Dark-flat:** a modification of the blob method that added a penalty for local luminance gradient in an attempt to reduce the influence of obliquely viewed walls.

The blob method was selected as the main method because a single darkest cell was unstable. In addition, when two separate dark regions were present, simply averaging all dark cells could produce a direction between the two regions, sometimes pointing toward a wall.

### 2.4 True wall-distance estimator

For each frame, the true distance to the colonic wall was calculated for each of the 576 directions by ray casting against the three-dimensional mesh using vtkOBBTree, with a maximum distance of 200 mm.

The same blob algorithm was then applied to the wall-distance values instead of image darkness. This produced a **distance-based lumen direction**, representing the direction that would be obtained if an ideal range sensor provided exact wall distances.

For computational efficiency, subsequent analyses used the rendered depth buffer. Agreement between the two distance methods was verified. The median absolute difference was 0.015 mm and the 90th percentile was 0.037 mm. Summary results for the distance-based direction were identical to one decimal place.

### 2.5 Geometric reference direction

A geometric reference was required to calculate angular error.

The local centreline tangent initially appeared to be the natural reference. However, the estimated lumen direction often corresponded to an open region substantially farther ahead. In a curved colon, the direction toward this distant open region can differ considerably from the local centreline tangent.

We therefore calculated the direction from the current position to a centreline point located a fixed arc length ahead.

Candidate distances of 10, 20, 40, 80, and 160 mm were examined. The main analysis was then performed using a fixed distance of **40 mm** for all frames, avoiding frame-by-frame selection of the most favourable reference.

## 3. Results

### 3.1 Brightness reflected lumen geometry only when distance attenuation was present

The distance-based estimator was identical under the two illumination conditions to six decimal places across all 342 frames. Thus, any difference between the two conditions was caused by illumination alone.

With point-source illumination, the median angular difference between the brightness-based and distance-based directions was **2.5°**. A difference greater than 30° occurred in **2.3%** of frames.

The within-frame rank correlation between brightness and true wall distance was negative in all **342 of 342 frames**, with a median Spearman correlation of **ρ = −0.770**. The relationship is shown in Figure 2.

**Figure 2.**
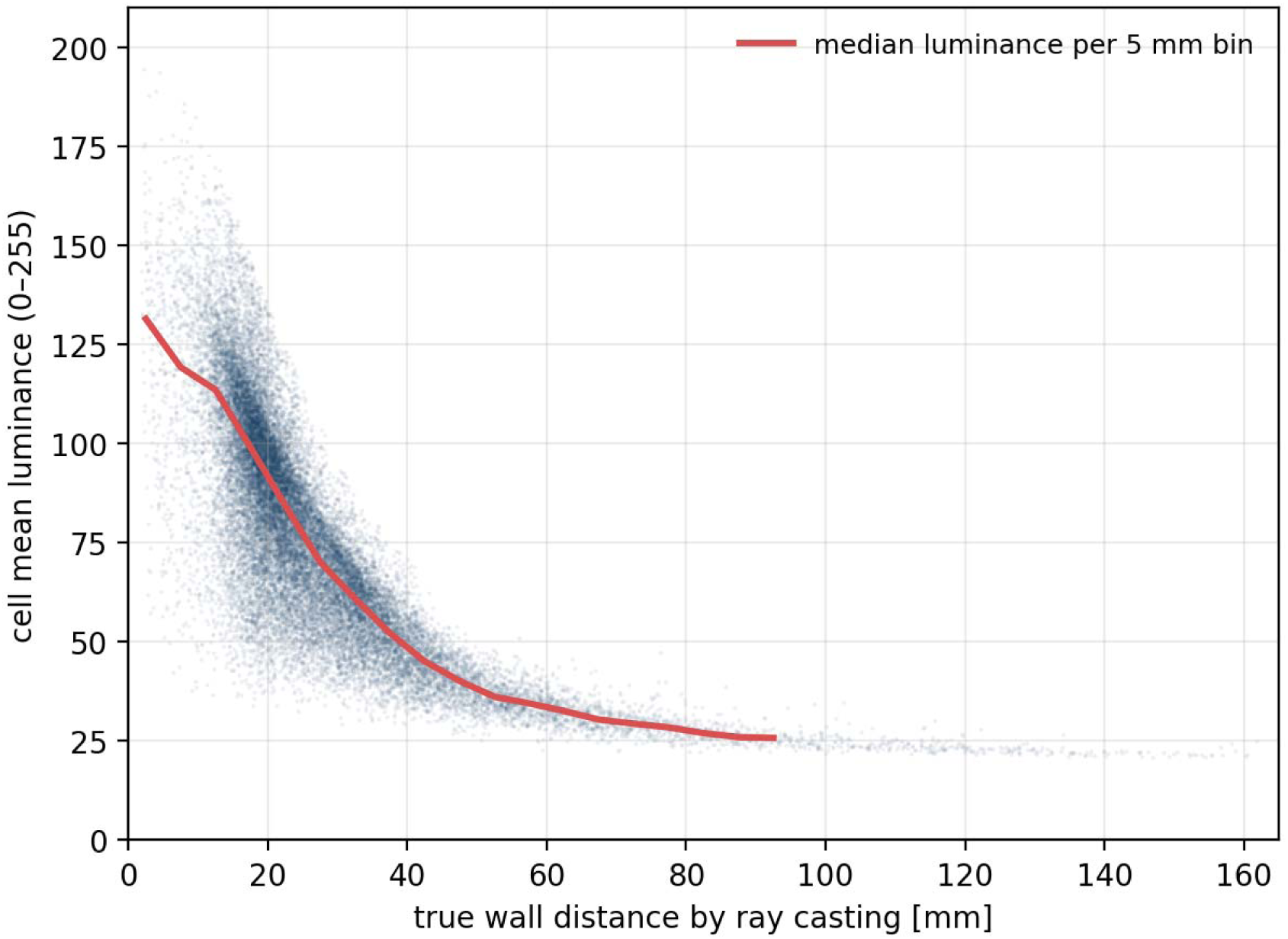
Cell luminance against true wall distance. Each point is one directional cell of one frame (28,386 cells pooled across the 342 analysed frames) under point-source illumination with inverse-square attenuation. The red line is the median luminance in 5 mm distance bins. The relationship is monotonic but one-to-many, because incidence angle also modulates radiance; brightness therefore orders directions by depth without measuring depth. The within-frame rank correlation was negative in every frame (median ρ = −0.770).

In contrast, directional illumination without distance attenuation did not provide a useful cue. The median angular difference between the brightness-based and distance-based directions increased to **43.1°**, and differences greater than 30° occurred in **76.5%** of frames. The median brightness–distance rank correlation was **ρ = +0.342**.

These results are summarised in Table 1. The directional information in image brightness depended strongly on distance-dependent illumination.

**Table 1.**
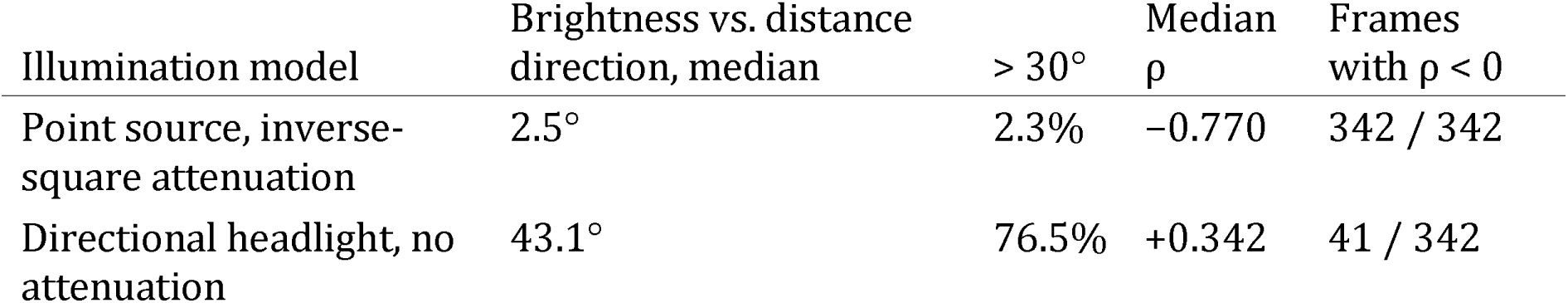
Effect of the illumination model. The distance-based estimator was identical under the two conditions; only the light differed.

| Illumination model | Brightness vs. distance direction, median | > 30° | Median $\rho$ | Frames with $\rho < 0$ |
| --- | --- | --- | --- | --- |
| Point source, inverse-square attenuation | 2.5° | 2.3% | -0.770 | 342 / 342 |
| Directional headlight, no attenuation | 43.1° | 76.5% | +0.342 | 41 / 342 |

ρ is the within-frame Spearman rank correlation between cell luminance and true wall distance.

### 3.2 Brightness-based and distance-based directions showed similar accuracy

Using the fixed 40 mm centreline chord as the geometric reference, the brightness-based blob estimator had a median angular error of **6.6°**, compared with **6.5°** for the true-distance estimator (Table 2).

**Table 2.** Angular error against a fixed geometric reference (the chord to the centreline point 40 mm ahead), 342 frames.

| Estimator | Median | 75th percentile | 90th percentile | Error > 30° |
| --- | --- | --- | --- | --- |
| Brightness (blob) | 6.6° | 14.5° | 24.8° | 7.3% |
| True wall distance (blob) | 6.5° | 13.3° | 24.3° | 6.7% |

The 75th-percentile errors were 14.5° and 13.3°, respectively, and the 90th-percentile errors were 24.8° and 24.3°. Errors greater than 30° occurred in 7.3% of frames for brightness and 6.7% for true distance.

When the two errors were subtracted frame by frame, the median difference attributable to the use of brightness rather than true distance was **−0.09°**.

Thus, for this particular task and virtual colon, replacing exact wall distance with image brightness produced very little loss of directional information.

When the reference chord length was allowed to vary among 10, 20, 40, 80, and 160 mm, 40 mm was selected in 62.2% of frames. At this length, the median angular error was 4.2°. The concentration of the selection at a single value, rather than a scatter across the candidates, indicates that approximately 40 mm is the range that the estimator was actually observing.

The choice of estimator also affected performance. The single-darkest-cell method was less stable than the blob method, particularly when multiple dark regions were visible.

### 3.3 Failure cases

Twenty-five frames had a brightness-based angular error greater than 30°. Their distribution along the colon is shown in Figure 3, and representative frames are shown in Figure 4.

**Figure 3.**
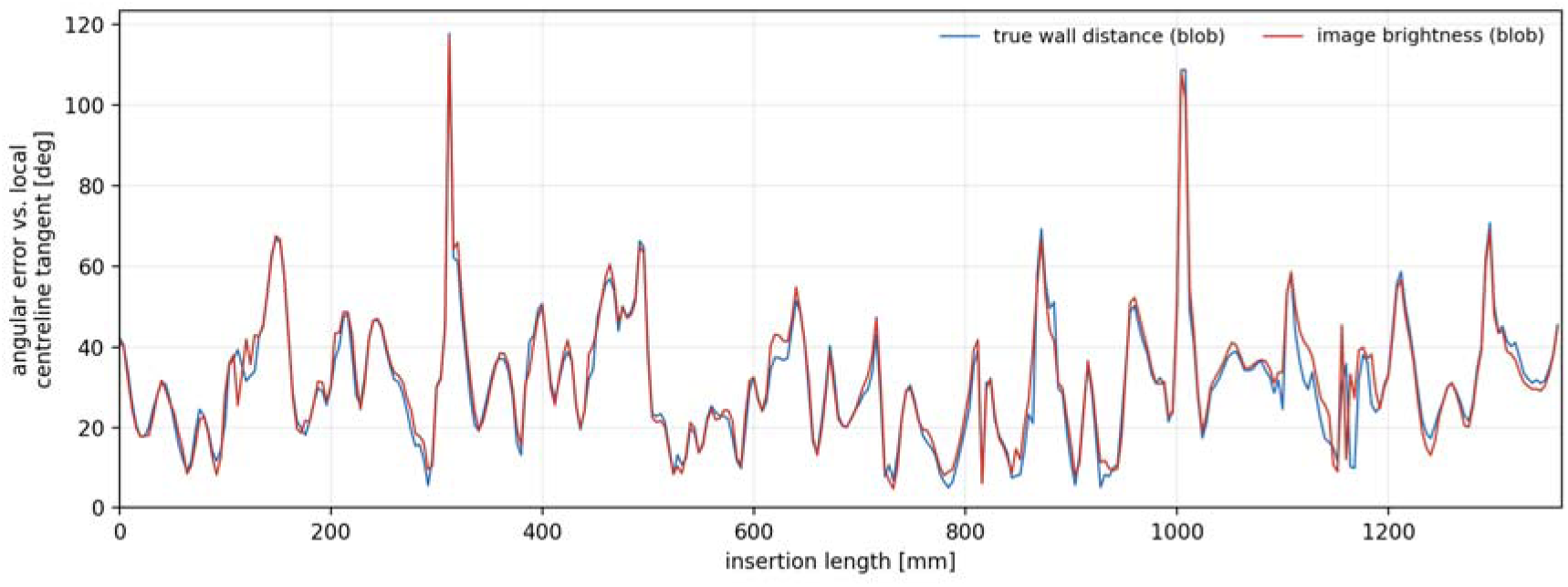
Angular error along the colon. Error of the brightness-based estimator (red) and of the true-distance estimator (blue), plotted against insertion length. Errors here are shown against the local centreline tangent, which is a stricter reference than the 40 mm chord used in Table 2 and is therefore systematically larger. The two traces move together, indicating that most of the residual error is common to both and is therefore attributable to the navigation rule rather than to the use of brightness.

**Figure 4.**
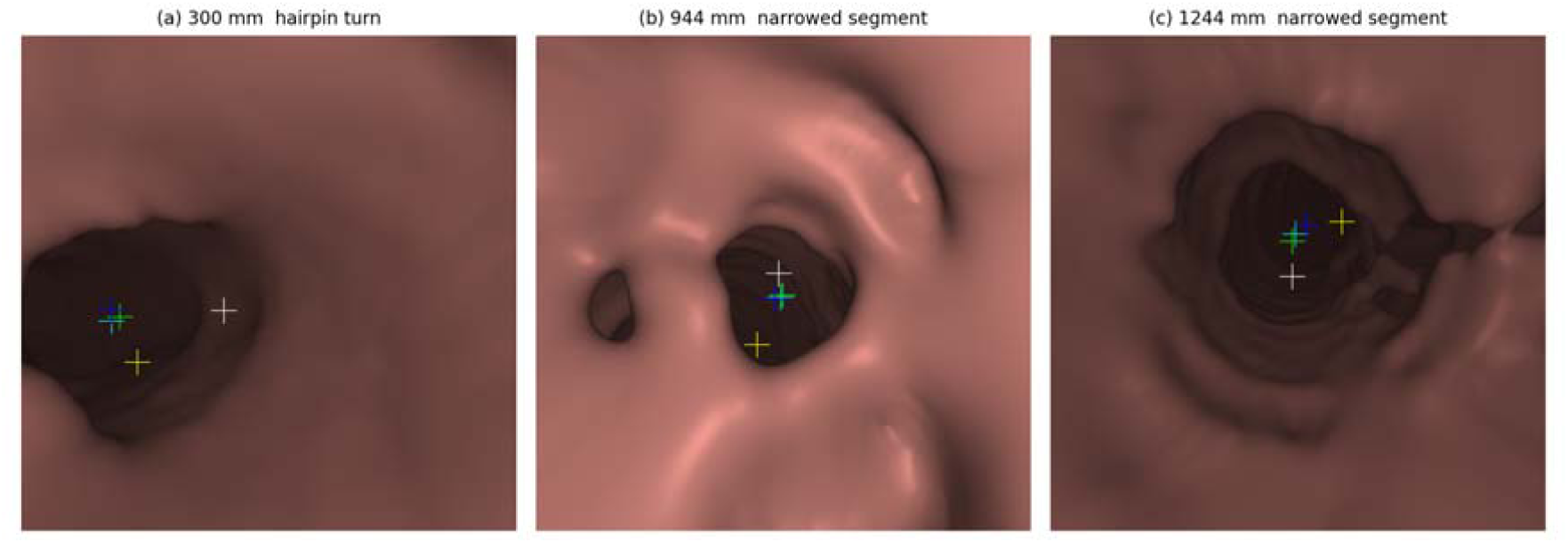
Representative frames at which the estimators disagreed with the geometric reference. White marker, local centreline tangent; cyan, brightness-based estimate; blue, distance-based estimate; yellow, single darkest cell. (a) A hairpin turn at 300 mm insertion: the brightness-based and distance-based estimates coincide, and both differ markedly from the reference, because the most open visible region is not the direction the instrument must take. (b, c) Narrowed segments at 944 mm and 1244 mm insertion. If such segments represent physiological collapse rather than fixed stenosis, they would distend under insufflation in vivo.

Several different mechanisms contributed to these failures (Table 3).

**Table 3.** Mechanisms underlying the 25 frames with a brightness-based angular error greater than 30°.

| Mechanism | Frames | Share of all frames | Distinguishing feature |
| --- | --- | --- | --- |
| Reference range mismatch | 12 | 3.5% | Error small when compared with a longer chord, typically 80 mm |
| Hairpin turn | 7 | 2.0% | Brightness-based and distance-based directions agree; both differ from the reference |
| Brightness-specific | 7 | 2.0% | Distance-based direction remains accurate; brightness-based direction does not |

In some frames, the estimated direction appeared incorrect when compared with the fixed 40 mm reference but agreed with a centreline point farther ahead, commonly at 80 mm. These cases occurred particularly in long, open segments in which the wall was visible up to 128 mm ahead.

Another group occurred at sharp hairpin turns. In these frames, the brightness-based and true-distance estimators agreed closely with each other, but both pointed in a direction very different from the 40 mm centreline reference. Thus, improved range sensing would not solve these failures. The problem was the navigation rule itself: the most open visible direction was not necessarily the direction required to negotiate a sharp turn.

A smaller group represented brightness-specific failures, in which the distance-based estimator remained close to the reference but the brightness-based estimator did not.

These observations distinguish two different problems: errors caused by brightness as a surrogate for distance, and errors caused by the more fundamental assumption that the instrument should always be directed toward the most open visible region.

## 4. Discussion

### 4.1 Main finding

This study tested one simple idea: whether the dark region of an endoscopic image can indicate the direction of the colonic lumen.

In the patient-derived virtual colon, the answer was largely yes.

With point-source illumination and distance attenuation, the direction calculated from image brightness differed from the direction calculated using exact wall distances by a median of only **2.5°**.

This does **not** mean that brightness provides an accurate measurement of wall distance. Brightness is affected by both distance and surface orientation, as well as by illumination and surface properties [11].

However, exact distance may not be necessary for steering.

For navigation, the important information may simply be:

**Which direction is more open?**

In the present virtual environment, brightness provided nearly the same answer to this question as exact wall-distance measurements.

### 4.2 A learning-free method for colonoscope steering

An important feature of the present method is that it does not require machine learning.

The algorithm consists of simple operations:

1. divide the image into directional cells;
2. identify the darkest cells;
3. find the largest connected dark region;
4. calculate its centre;
5. steer the endoscope toward that direction.

No training dataset is required, and no learned parameters are used.

This distinction may be useful for automated colonoscopy. Recent systems have applied convolutional networks, reinforcement learning and end-to-end visuomotor control to the same task [1,2,16]. If a useful navigation signal can be obtained directly from the physics of endoscopic illumination, there may be little advantage in asking a machine-learning model to learn the same relationship from a large dataset.

The present study therefore suggests a simple control sequence:

**image brightness ⍰ lumen direction ⍰ calculated angulation**

rather than:

**image ⍰ machine-learning model ⍰ predicted action**.

This does not imply that machine learning has no role in future automated colonoscopy. Machine learning may become useful for difficult anatomical situations, image artefacts, depth estimation, lesion recognition, or more complex control. However, the basic task of maintaining the tip toward an open lumen may be achievable by deterministic calculation.

A simple method also has practical advantages. Its behaviour can be inspected directly, its failure conditions can be identified, and the reason for each steering command can be understood. These properties are relevant to regulatory evaluation of increasing levels of device autonomy [18].

### 4.3 Automatic angulation release

Finding the lumen direction is only part of colonoscope control.

During manual colonoscopy, the operator applies angulation to negotiate a bend. After the tip has passed the bend, the angulation should be reduced. Maintaining excessive angulation while continuing to advance the instrument can increase contact with the wall and make further insertion difficult.

We therefore propose a second computational function: **automatic angulation release**, or *auto-unwind*.

This does not require the system to identify a specific anatomical flexure or to decide explicitly that a turn has been completed.

Instead, the same lumen-direction signal can be used continuously.

When the estimated lumen direction is away from the optical axis, the system applies the angulation required to follow it. As the lumen returns toward the optical axis, the applied angulation is gradually relaxed toward neutral. If relaxation causes the lumen direction to move away from the optical axis again, relaxation stops.

The control objective can therefore be stated simply:

**Use the minimum angulation necessary to keep the lumen in the desired direction.**

This changes the control problem from a sequence of discrete decisions into continuous regulation.

In clinical terms, the system moves from **maximum bend toward minimum necessary bend** as soon as the geometry permits.

Like the brightness-based direction estimator, this control rule can be implemented by calculation without machine learning.

### 4.4 A semi-autonomous rather than fully autonomous colonoscope

The present findings do not require a fully autonomous colonoscope.

We propose a simpler division of labour between the computer and the endoscopist.

The **computer** performs:

- fine up/down and left/right tip steering;
- maintenance of the lumen direction near the optical axis; and
- automatic release of unnecessary angulation.

The **operator** performs:

- insertion;
- withdrawal;
- major shaft rotation; and
- loop reduction.

In ordinary insertion, the operator could therefore advance the colonoscope slowly while the system continuously directs the tip toward the lumen and reduces unnecessary angulation.

This arrangement may be described as **semi-autonomous colonoscopy**. It leaves the frequent, repetitive task of fine tip steering to the computer while preserving direct human control over advancement and the more complex mechanical problem of loop management. In existing frameworks for medical device autonomy, this corresponds to task-level rather than conditional or high autonomy [18]. Comparable divisions of labour have been implemented in robotised flexible endoscopes with automated lumen centralisation, in which the automatic function was engaged intermittently and at the operator’s discretion [14,15].

### 4.5 Loop reduction can remain a manual procedure

Loop formation is fundamentally different from lumen-direction control.

The lumen direction is mainly a local problem at the distal tip. In contrast, loop formation involves the shape of a much longer section of the colonoscope shaft and its interaction with the colon [19,20].

A fully autonomous system might therefore require additional information about shaft configuration.

For the semi-autonomous system proposed here, this is unnecessary.

When advancement becomes ineffective because a loop has formed, the operator can stop automatic advancement, withdraw the colonoscope, and manually reduce the loop. As the loop becomes smaller, the operator can rotate the shaft and continue withdrawal or straightening until a more direct configuration is obtained. These are established manual manoeuvres [20].

The proposed system therefore does **not** need to identify the exact type of loop or automatically determine the correct direction of shaft rotation.

This is an intentional division of labour:

**The computer controls local tip direction; the endoscopist manages global shaft configuration.**

An electromagnetic colonoscope imaging system, when available, could provide additional confirmation of shaft configuration and straightening [19]. However, such imaging is not essential to the basic concept proposed here.

Importantly, loop reduction was not tested in the present simulation because the current virtual colonoscope does not reproduce deformable shaft mechanics. It should therefore be regarded as a proposed component of the future semi-autonomous system rather than a result of this study.

### 4.6 Why exact depth may not be necessary

Much work on automated endoscopic navigation focuses on estimating depth or reconstructing three-dimensional geometry [3,4,5,21].

These remain important technologies. They may ultimately provide information about tip position, trajectory, wall distance, and the shape of the surrounding lumen.

However, the present experiment suggests that **metric depth and steering direction should be considered separately**.

A system may not know that the wall is exactly 18 mm away and still know that the right side is more open than the left.

For basic tip steering, this relative directional information may be sufficient.

This distinction could simplify the first generation of semi-autonomous colonoscope systems. More sophisticated sensing could subsequently be added for tasks that genuinely require metric information, such as three-dimensional tip tracking or reconstruction of the colonoscope trajectory.

### 4.7 Sharp turns reveal a limitation of the simple navigation rule

The failure analysis also identified an important limitation.

At several hairpin turns, the brightness-based and true-distance estimators agreed closely with each other, but both pointed in the wrong direction relative to the future centreline (Figure 4a).

This is important because it shows that the problem cannot be solved simply by improving distance measurement.

At a sharp bend, the most open visible region may lie in the direction from which the colonoscope has just arrived, while the correct path requires turning toward a smaller and less obvious opening.

Thus,

**“steer toward the most open direction” is useful, but it is not a complete navigation strategy.**

These situations are likely to be an important target for future work.

They may be handled by adding information from previous frames, insertion direction, recent tip trajectory, or a simple state-based control rule. More complex methods, including machine learning, could also be considered if deterministic rules are insufficient.

The present simulator provides an environment in which these alternatives can be tested directly.

### 4.8 Collapsed segments and stenosis

Some regions of the CT-derived colon were very narrow. In a static virtual colon, such regions may be impossible for a realistically sized virtual colonoscope to traverse.

In vivo, however, a physiologically collapsed segment may open during insufflation. A fixed pathological stenosis may not.

Because the present model was derived from a single static CT geometry, it cannot reproduce this distinction directly.

Some brightness-specific failures occurred when the modelled wall was within 2.3 mm of the virtual tip (Figure 4b, 4c). If these regions represent physiological collapse rather than fixed narrowing, these failures may partly reflect the limitations of the static phantom, and the brightness-specific failure rate of 2.0% should be regarded as an upper bound.

CT colonography obtained in more than one patient position may help distinguish deformable collapse from fixed narrowing [17] and may allow future versions of the phantom to represent these regions more realistically.

### 4.9 Limitations

This study has several important limitations.

First, only one patient-derived colon was studied. Anatomical variation between patients was therefore not evaluated.

Second, the rendering model was idealised. A real colonoscope has light guides that are not perfectly co-located with the camera. Real images are also affected by automatic gain control, vignetting, gamma correction, specular reflections, mucus, fluid, bubbles, stool, and other material. These factors may alter the relationship between brightness and lumen direction.

Third, the virtual colon was static. It did not reproduce insufflation, peristalsis, deformation caused by the colonoscope, or movement of the colon itself.

Fourth, the colonoscope shaft was not mechanically simulated. Loop formation and loop reduction therefore could not be tested. The proposed human–computer division of labour for loop management remains a future concept.

Fifth, the present experiment was performed with the camera located on the centreline and initially directed along the lumen. During actual semi-autonomous navigation, the tip will move away from the centreline and may view the wall at more oblique angles. Closed-loop testing is therefore necessary.

Finally, the present experiment tested only **direction estimation**. It did not test whether a virtual colonoscope controlled by this estimator can successfully reach the caecum. That is the next step.

## 5. Future work

The immediate next experiment is to convert the present direction estimator into a closed-loop controller in the virtual colon.

The virtual colonoscope will be advanced while the system automatically controls up/down and left/right angulation according to the brightness-derived lumen direction.

Three functions will be evaluated:

**1. automatic lumen-directed steering;**
**2. automatic release of unnecessary angulation; and**
**3. real-time recording of the three-dimensional tip trajectory.**

The operator will retain control of insertion and withdrawal and, in future mechanical simulations or physical systems, loop reduction.

Because the virtual environment provides exact wall geometry and exact tip position, it also allows the brightness-based controller to be compared directly with controllers using perfect geometric information.

This provides a stepwise path from a simple computational observation to a semi-autonomous colonoscopy system without requiring machine learning at the initial stage.

## 6. Conclusion

In a virtual colon reconstructed from patient CT data, lumen direction estimated from image brightness closely matched the direction estimated from exact wall distances. With point-source illumination and distance attenuation, the median difference between the two directions was **2.5°**. Against a fixed 40 mm centreline reference, median errors were **6.6° for brightness and 6.5° for exact distance**.

These results do not show that brightness can measure wall distance. Rather, they suggest that **exact distance may not be necessary for the simpler task of determining which direction the lumen continues**.

On this basis, we propose a learning-free approach to semi-autonomous colonoscopy in which image brightness is converted directly into a lumen direction by calculation. The system performs fine tip steering and automatically reduces unnecessary angulation, while the endoscopist retains control of insertion, withdrawal, shaft rotation, and loop reduction.

This division of labour may provide a practical intermediate step between conventional manual colonoscopy and fully autonomous colonoscopy.

## Ethics statement

The Institutional Review Board of Kojima Family Clinic gave ethical approval for this study. The CT colonography data used to construct the virtual colon were the author’s own pre-existing medical imaging data and were originally acquired for clinical purposes, not specifically for this study. No data from other patients or research participants were used. The images were de-identified before computational analysis.

## Data and code availability

The analysis code (dark_probe.py, autopilot.py) and the virtual colonoscopy simulator are being prepared for public release and will be made available in a public repository. The original CT data are the author’s personal medical imaging data and will not be publicly shared.

## Data Availability

The data generated in this study are contained within the manuscript. The analysis code and virtual colonoscopy simulator are being prepared for public release. The original CT colonography data are the author's personal medical imaging data and will not be publicly shared.

## Acknowledgements

The author used Claude (Anthropic) and ChatGPT (OpenAI) to assist with English-language editing and manuscript preparation. All study design, computational experiments, data analysis, interpretation of the results, and final decisions regarding the manuscript were performed by and remain the responsibility of the author.

